# Preferences and Acceptability for Long-Acting PrEP Agents Among Pregnant and Postpartum Women with Experience Using Daily Oral PrEP in South Africa and Kenya

**DOI:** 10.1101/2022.10.29.22281701

**Authors:** Nafisa J. Wara, Rufaro Mvududu, Mary M. Marwa, Laurén Gómez, Nyiko Mashele, Catherine Orrell, Corrina Moucheraud, John Kinuthia, Grace John-Stewart, Landon Myer, Risa Hoffman, Jillian Pintye, Dvora L. Joseph Davey

## Abstract

**Introduction:** Long-acting pre-exposure prophylaxis (PrEP) options could overcome some barriers to oral PrEP persistence during pregnancy and postpartum. We evaluated long-acting PrEP preferences among oral PrEP-experienced pregnant and postpartum women in South Africa and Kenya, two countries with high coverage of oral PrEP and with pending regulatory approvals for long-acting injectable cabotegravir and the dapivirine vaginal ring (approved in South Africa, under review in Kenya).

**Methods:** From September 2021 to February 2022, we surveyed pregnant and postpartum women enrolled in oral PrEP studies in South Africa and Kenya. We evaluated oral PrEP attitudes and preferences for existing and future long-acting PrEP methods.

**Results:** We surveyed 190 women in South Africa (67% postpartum; median age 27 years [IQR 22-32]) and 204 women in Kenya (79% postpartum; median age 29 years [IQR 25-33]). 75% of participants reported oral PrEP use within the last 30 days. Overall, 49% of participants reported negative oral PrEP attributes, including side effects (21% South Africa, 30% Kenya) and pill burden (20% South Africa, 25% Kenya). Preferred PrEP attributes included long-acting method, effectiveness, safety while pregnant and breastfeeding, and free medication. Most participants (75%, South Africa and Kenya) preferred a potential long-acting injectable over oral PrEP, most frequently for longer duration of effectiveness in South Africa (87% South Africa, 42% Kenya) versus discretion in Kenya (5% South Africa, 49% Kenya). 87% of participants preferred oral PrEP over a potential long-acting vaginal ring, mostly due to concern about possible discomfort with vaginal insertion (82% South Africa, 48% Kenya). Significant predictors of long-acting PrEP preference included past use of injectable contraceptive (aOR 2.48, 95% CI: 1.34, 4.57), disliking at least one oral PrEP attribute (aOR 1.72, 95% CI: 1.05, 2.80), and preferring infrequent PrEP use (aOR 1.58, 95% CI: 0.94, 2.65).

**Conclusions:** Oral PrEP-experienced pregnant and postpartum women expressed a theoretical preference for long-acting injectable PrEP over other modalities, demonstrating potential acceptability among a key population who must be at the forefront of injectable PrEP rollout. Reasons for PrEP preferences differed by country, emphasizing the importance of increasing context-specific options and choice of PrEP modalities for pregnant and postpartum women.

## Introduction

HIV incidence remains high among cisgender women of reproductive age in South Africa and Kenya, including during pregnancy and postpartum[1-4]. Pregnant women without HIV are at elevated risk of acquisition due to structural and sociocultural factors (e.g., poverty, gender inequity) that may result in high-risk scenarios, including not knowing the HIV status of their partner(s), engaging in sexual activity without condoms, and having multiple sex partners[5-8]. Furthermore, the risk of vertical HIV transmission is elevated during pregnancy and breastfeeding due to acute maternal HIV infection[9,10]. A concerted effort is needed to reduce HIV infections among pregnant and breastfeeding individuals, and their infants.

Daily oral tenofovir disoproxil fumarate/emtricitabine (TDF/FTC) pre-exposure prophylaxis (PrEP) for HIV prevention is being rapidly scaled in South Africa and Kenya, and these two countries have the highest and second highest number of PrEP initiations globally[11]. Studies have reported high rates of oral PrEP uptake among pregnant and postpartum women in South Africa and Kenya[12,13]. However, the delivery of daily oral PrEP among pregnant and postpartum women is challenged by insufficient integration of PrEP provision and counseling into existing antenatal and postpartum systems[14]. Furthermore, self-reported adherence on oral PrEP (e.g., taking PrEP daily) was low among pregnant and postpartum women in Kenya, and limited adherence was identified among pregnant and postpartum women taking oral PrEP in South Africa through dried blood spot analysis[13,15]. Substantial barriers to oral PrEP adherence exist in these populations, including pill burden, stigma and limited disclosure of PrEP use, and financial and logistical barriers to accessing a clinic for PrEP[16-18]. Strategies to overcome barriers to PrEP adherence are urgently needed.

Long-acting modalities, such as injectable cabotegravir (CAB-LA) and the dapivirine vaginal ring, may improve PrEP initiation and persistence among pregnant and postpartum women. CAB-LA, an intramuscular injection administered every eight weeks, is superior to oral PrEP, reducing risk of HIV infection by 88% compared to daily oral TDF/FTC among cisgender women in Sub-Saharan Africa[19]. Preliminary data show that CAB-LA is well tolerated during pregnancy and has a similar pharmacokinetic profile to its use by nonpregnant women[20,21]. CAB-LA received regulatory approval in the United States in December 2021 and is pending regulatory approval in South Africa, Kenya, and other countries in Southern Africa[11]. The dapivirine ring, inserted vaginally and replaced every four weeks, has been shown to reduce the risk of HIV infection by approximately 30% and is not associated with adverse pregnancy or infant outcomes[22]. The dapivirine ring received regulatory approval in South Africa in March 2022, and is pending approval in Kenya[11]. These developments highlight the urgent need to understand potential facilitators and barriers to uptake and persistence on these novel PrEP methods among pregnant and postpartum women. Our study aims to assess preferences for and perceptions of long-acting PrEP methods among oral PrEP-experienced pregnant and postpartum women in Kenya and South Africa.

## Methods

### Study participants

We conducted surveys with participants enrolled in an observational cohort study assessing daily oral PrEP initiation and persistence among pregnant and breastfeeding women (PBFW) in Cape Town, South Africa (PrEP-PP) and an observational extension cohort of a cluster randomized trial offering daily oral PrEP among PBFW in Western Kenya (PrIMA-X)[23,24]. Eligibility criteria for PrEP-PP included: ≥16 years old, confirmed HIV-negative serostatus by a 4^th^ generation antigen/antibody combination HIV test (Abbott), intention to stay in Cape Town through the postpartum period, and no contraindications to PrEP use. Eligibility criteria for PrIMA-X included: ≥15 years old, confirmed HIV and TB negative, intention to reside in the area for at least one year postpartum, and plans to receive postnatal and infant care at the study facility. Enrolment criteria for this subsample included: currently using or having previously used daily oral PrEP; currently pregnant or ≤9 months postpartum; and enrolment in PrEP-PP or PrIMA-X.

### Data collection

Between September 2021 and February 2022, trained study staff fluent in English and either isiXhosa, Kiswahili, or Luo approached women attending PrEP-PP or PrIMA-X follow-up visits to introduce the study. Study staff then screened participants for study eligibility, obtained written informed consent in English or the participant’s local language (isiXhosa, Kiswahili, or Luo), and administered the survey to eligible consenting participants. Study staff asked participants survey questions and recorded responses on a tablet. The survey took 30-40 minutes and was completed on REDCap, a secure web-based platform[25]. Participants received 120 Rand in South Africa (∼$7 USD) or KSh 300 in Kenya (∼$3 USD) for their participation in the study as well as transportation expenses.

### Survey Measures

#### Participant Sociodemographic Characteristics

We collected (a) basic demographic data (including age, obstetric history, education level, employment, number of sexual partners), (b) HIV risk perception (response to “How would you describe your chances of getting HIV in the next year?” from “no chance at all” to “great chance”), (c) current alcohol use adapted from the Alcohol Use Disorders Identification Test[26,27], (d) PrEP adherence based on 30-day recall (i.e. responding “yes” or “no” to taking PrEP within the past 30 days), (e) clinic access (including transportation method, total travel time, and total transportation cost), and (f) previous use of any contraceptive methods. We also assessed PrEP stigma using a seven-item scale derived from existing literature in which participants responded to statements describing experiences of PrEP stigma via a 5-point Likert scale ranging 0 = “strongly disagree” to 4 = “strongly agree”[28,29].

#### Current and Future PrEP Preferences

We assessed perceptions of daily oral PrEP by asking participants what they like and dislike about oral PrEP. We described methods of long-acting HIV prevention currently pending regulatory approval or in development, and asked participants about preferences regarding HIV prevention modalities that may be available in the future. We adapted a list of PrEP characteristics from a discrete choice experiment of HIV prevention methods assessed within the Quatro Study[30]. We asked participants to rank the top three most important characteristics of a potential HIV prevention product, the top three most important access-related characteristics, and how frequently they would theoretically use an HIV prevention method.

#### Long-acting PrEP Preferences

We assessed preferences regarding injectable PrEP and the vaginal ring by asking participants whether they would prefer to switch to the long-acting method or remain on oral PrEP. We then assessed reasons for preferring the long-acting method or oral PrEP.

### Statistical analyses

We used descriptive statistics (median, interquartile range [IQR], frequency) to report participant responses, and used Chi-square, Fischer’s Exact, and Wilcoxon rank sum to compare responses between countries. We used univariate and multivariable logistic regression models to assess predictors of preferring a long-acting PrEP method (injection or ring) over oral PrEP (among those with this preference). We adjusted for *a priori* potential confounders (maternal age and country) in the multivariable models. Statistical analyses were conducted with STATA v.17[31].

### Ethics

The PrEP-PP study was approved by the Human Research Ethics Committee of the University of Cape Town Faculty of Health Sciences (#297/2018) and by the University of California, Los Angeles Institutional Review Board (IRB#18-001622). The PrIMA-X study was approved by the Kenyatta National Hospital-University of Nairobi Ethics and Research Committee (P73/02/2017) and by the University of Washington Human Subjects Division (STUDY00000438).

## Results

Overall, 394 women were enrolled in the study, 190 women from the South African cohort and 204 from the Kenyan cohort. Median ages were 27 (IQR=22-32) and 29 (IQR=25-33) respectively (**Table 1**). Overall, 33% of South African participants were pregnant (n=63) while 21% of Kenyan participants were pregnant (n=42), with the remaining in both groups postpartum. Most participants completed some or all secondary school education (total n=326, 83%) although secondary education was more common in South Africa (93%, Kenya 74%, p<0.01). Unemployment was more common in Kenya (Kenya 87%, South Africa 72%, p<0.01). Almost all (95%) women reported having at least one current sexual partner (n=373). Most participants reported that they took at least one dose of PrEP over the previous 30 days at the time of the survey (South Africa 82%, Kenya 68%, p<0.001), and median time on PrEP among current PrEP users was 337 days (IQR=263-420) and 308 days (IQR=114-442) among South African and Kenyan participants, respectively.

**Table 1:** Demographics and health characteristics of pregnant and postpartum women with experience taking oral PrEP, South Africa and Kenya, September 2021 – February 2022 (N=394 women)

|  | **Overall**<br>**(N=394, %)** | **South Africa**<br>**(n=190, %)** | **Kenya**<br>**(n=204, %)** | **p-value** |
| --- | --- | --- | --- | --- |
| Age (median, IQR) | 28 [24-32] | 27 [22-32] | 29 [25-33] | <0.01 |
| Pregnant (n, %) | 105 (27) | 63 (33) | 42 (21) | 0.01 |
| Postpartum (n, %) | 289 (73) | 127 (67) | 162 (79) | 0.01 |
| Days on oral PrEP (median, IQR) | 335 [168-420] | 337 [263-420] | 308 [114-442] | 0.04 |
| Last grade completed (n, %) |  |  |  |  |
| Primary school (Grades 1-6) | 28 (7) | 2 (1) | 26 (13) | <0.01 |
| Some secondary school (Grades 7-11) | 236 (60) | 106 (56) | 130 (64) |  |
| Completed secondary school | 90 (23) | 70 (37) | 20 (9) |  |
| Some tertiary | 16 (4) | 8 (4) | 8 (4) |  |
| Completed tertiary | 24 (6) | 4 (2) | 20 (10) |  |
| Currently employed (formally or informally)? (n, %) |  |  |  |  |
| No | 313 (79) | 136 (72) | 177 (87) | <0.01 |
| Yes | 80 (20) | 54 (28) | 26 (13) |  |
| Prefer not to answer | 1 (1) | 0 (0) | 1 (1) |  |
| Currently have at least one sexual partner (n, %) |  |  |  |  |
| No | 21 (5) | 15 (8) | 6 (3) | 0.03 |
| Yes | 373 (95) | 175 (92) | 198 (97) |  |
| HIV Risk Perception<sup>†</sup> (n, %) |  |  |  |  |
| No risk at all | 121/380 (32) | 84/176 (48) | 37 (18) | <0.01 |
| Small chance | 145/380 (44) | 56/176 (32) | 89 (44) |  |
| Moderate chance | 89/380 (34) | 20/176 (11) | 69 (34) |  |
| Great chance | 23/380 (6) | 16/176 (9) | 7 (3) |  |
| Prefer not to answer | 2/380 (1) | 0/176 (0) | 2 (1) |  |
| Current alcohol use (n, %) |  |  |  |  |
| Never | 363/380 (96) | 164/176 (93) | 199 (98) | 0.31 |
| Monthly or less | 11/380 (3) | 8/176 (5) | 3 (2) |  |
| 2-4 times a month | 5/380 (1) | 3/176 (2) | 2 (1) |  |
| 2-3 times a week | 1/380 (1) | 1/176 (1) | 0 (0) |  |
| 4 or more times a week | 0 (0) | 0 (0) | 0 (0) |  |
| HIV PrEP Stigma<sup>‡</sup> (n, %) |  |  |  |  |
| Endorsed 0 forms of stigma | 251/380 (66) | 128/176 (73) | 123 (60) | 0.01 |
| Endorsed >1 form of stigma | 129/380 (34) | 48/176 (27) | 81 (40) |  |
| Any oral PrEP use over past 30 days (n, %) |  |  |  |  |
| No | 100 (25) | 34 (18) | 66 (32) | <0.01 |
| Yes | 294 (75) | 156 (82) | 138 (68) |  |
| Clinic transportation (n, %) |  |  |  |  |
| Walking | 31 (8) | 16 (8) | 15 (7) | <0.01 |
| Taxi/Minibus | 206 (52) | 172 (91) | 34 (17) |  |
| Private car | 3 (1) | 2 (1) | 1 (1) |  |
| Motorbike | 152 (38) | 0 (0) | 152 (75) |  |
| Other | 2 (1) | 0 (0) | 2 (1) |  |
| Total travel time to clinic and return (minutes, median, IQR) | 40 [30-60] | 30 [20.0-40.0] | 55 [31.0-60.0] | <0.01 |
| Cost to travel to clinic and return (USD, median, IQR) | 1.32 [1.31-1.74] | 1.32 [1.32-1.32] | 1.75 [0.88-2.64] | 0.02 |

<sup>†</sup> Participants were asked “How would you describe your chances of getting HIV in the next year?” with the option to select “No risk at all,” “Small chance,” “Moderate chance,” “Great chance,” or “Prefer not to answer.”
<sup>‡</sup> Participants were asked to respond on a 5-point Likert scale from “Strongly Disagree” to “Strongly Agree” to 7 statements on potential stigma experienced regarding PrEP use (i.e. “I feel ashamed of using PrEP,” “I feel embarrassed about using PrEP”).

Have you ever used any of the following family planning methods? (n, %)
|  |  |  |  |  |
| --- | --- | --- | --- | --- |
| Male condom | 217 (55) | 171 (90) | 46 (23) | <0.01 |
| Oral contraceptive pill | 98 (25) | 42 (22) | 56 (27) | 0.22 |
| Injectable contraceptive | 312 (79) | 178 (94) | 134 (66) | <0.01 |
| Contraceptive implant | 181 (46) | 70 (37) | 111 (54) | <0.01 |
| Female condom | 8 (2) | 6 (3) | 2 (1) | 0.97 |
| Vaginal contraceptive ring | 2 (1) | 0 (0) | 2 (1) | 0.27 |
| IUD/loop | 6 (2) | 3 (2) | 3 (2) | 0.69 |
| Other (tubal ligation) | 2 (1) | 0 (0) | 2 (1) | 0.27 |
| None | 11 (3) | 1 (1) | 9 (4) | 0.01 |

At entry into the parent study, 52% of South African women and 81% of Kenyan women reported any perceived risk of HIV acquisition (p<0.01). More than half endorsed no forms of PrEP stigma (66%, n=251/380), although more women in Kenya endorsed at least one form of PrEP stigma (Kenya 40%, South Africa 27%, p=0.01).

At the time of the survey, participants’ median travel time to and from the clinic was 30 minutes (IQR=20-40) in South Africa and 55 minutes (IQR=31-60) in Kenya (p<0.01), and median cost to travel to and from the clinic was USD $1.32 (IQR=1.32-1.32) and $1.75 (IQR=0.88-2.64) respectively (p=0.01. When asked about any previous experience with contraceptives, women frequently described use of injectable contraceptives (South Africa 94% vs Kenya 66%, p<0.01), followed in South Africa by use of male condoms (n=171, 90%) and contraceptive implants (n=70, 37%). Among women in Kenya, injectable contraceptive was most common (n=134, 66%), followed by the contraceptive implant (n=111, 54%) and then male condoms (n=46, 23%).

### Experiences with Oral PrEP

Almost all participants (n=386, 98%) reported efficacy in HIV prevention as a positive characteristic of daily oral PrEP, followed by daily oral PrEP having few or no side effects (n=61, 15%) and being easy to use (n=27, 7%) (**Table 2**). In addition, 10 (5%) participants in Kenya (compared to none in South Africa) liked that oral PrEP was discreet and 5 (2%) participants in South Africa (compared to none in Kenya) reported oral PrEP’s safety during breastfeeding as a positive characteristic. Forty-two percent of women in South Africa (n=79) and 56% in Kenya (n=114, p<0.01) disliked at least one attribute about daily oral PrEP. The most frequently reported dislikes included side effects (South Africa 21%, Kenya 30%, p=0.03), daily use (South Africa 20%, Kenya 25%, p=0.20), and taking it orally (South Africa 6%, Kenya 11%, p=0.08).

**Table 2:** Likes and dislikes regarding daily oral PrEP in pregnant and postpartum women with experience taking oral PrEP, South Africa and Kenya, September 2021 – February 2022.

| **Oral PrEP likes** | **Overall**<br>**(N=394)** | **South Africa**<br>**(n=190)** | **Kenya**<br>**(n=204)** | ***p*-value** |
| --- | --- | --- | --- | --- |
| HIV prevention | 386 (98) | 186 (98) | 200 (98) | 0.92 |
| PrEP has few/no side effects | 61 (15) | 35 (18) | 26 (13) | 0.12 |
| Ease of use | 27 (7) | 16 (8) | 11 (5) | 0.23 |
| No interruption of sex (as is needed for condoms) | 16 (4) | 5 (3) | 11 (5) | 0.17 |
| Easy to hide | 10 (3) | 0 (0) | 10 (5) | <0.01 |
| Take it daily | 8 (2) | 5 (3) | 3 (2) | 0.88 |
| Taken orally | 7 (2) | 5 (3) | 2 (1) | 0.95 |
| PrEP is safe for the baby | 5 (1) | 5 (3) | 0 (0) | 0.03 |
| Other (e.g., overall safety, increased appetite, peaceful sleep) | 4 (1) | 4 (2) | 0 (0) | 0.05 |
| Nothing | 1 (1) | 0 (0) | 1 (1) | 0.52 |
| **Oral PrEP dislikes** | **Overall**<br>**(N=394)** | **South Africa**<br>**(n=190)** | **Kenya**<br>**(n=204)** | ***p*-value** |
| Side effects | 100 (25) | 39 (21) | 61 (30) | 0.03 |
| Must take it daily | 90 (23) | 38 (20) | 52 (25) | 0.20 |
| Must take orally | 35 (9) | 12 (6) | 23 (11) | 0.08 |
| Pill size is too big | 12 (3) | 9 (5) | 3 (2) | 0.99 |
| No STI prevention | 10 (3) | 4 (2) | 6 (3) | 0.42 |
| Not discreet | 9 (2) | 0 (0) | 9 (4) | <0.01 |
| Other (e.g., general dislike, pill taste, pill smell) | 3 (1) | 2 (1) | 1 (1) | 0.90 |
| Nothing | 201 (51) | 111 (58) | 90 (44) | <0.01 |

### Future PrEP Preferences

When asked to rank characteristics of a potential PrEP product that may be available to them in the future, 52% of participants (n=203) ranked effectiveness at preventing HIV as the most important, followed by the ability to have a healthy pregnancy (n=47, 12%), frequency of use (n=39, 10%), tolerability (n=39, 10%), and the ability to breastfeed and have a healthy baby (n=26, 7%). However, there were country-specific differences in the ranking of several attributes: more women in Kenya ranked HIV prevention highest (78%, South Africa 23%, p<0.01), more women in South Africa ranked having a healthy pregnancy (19%, Kenya 5%, p<0.01), frequency of use (18%, Kenya 2%, p<0.01), side effects (16%, Kenya 4%, p<0.01), and privacy (6%, Kenya 1%, p=0.01) as most important (**Table 3**). When asked to rank access-related characteristics of a potential PrEP product, 52% of participants ranked medication being free as the most important, followed by ease of the process by which the product is obtained, location, and total time it takes to get the product. When asked to rank preferred frequency of PrEP use, participants most frequently preferred once a year (n=123, 31%), followed by once per month (n=62, 16%), once every 2-3 months (n=59, 15%), before sex (n=53, 13%), every day (n=46, 12%), once every 6 months (n=45, 11%), and after sex (n=6, 2%). Kenyan women preferred PrEP use once every 6 months, or once every 2-3 months, whereas South African women preferred a product that could be used once a year (p<0.01).

**Table 3:** Ranked characteristics of potential HIV prevention products among pregnant and postpartum women with experience taking oral PrEP, South Africa and Kenya, September 2021 – February 2022.

| **Most important characteristic of a potential HIV prevention product (ranked #1)** | **Overall (N=394)** | **South Africa (n=190)** | **Kenya (n=204)** | **p-value** |
| --- | --- | --- | --- | --- |
| Effective at preventing HIV | 203 (52) | 44 (23) | 159 (78) | <0.01 |
| Healthy pregnancy | 47 (12) | 36 (19) | 11 (5) | <0.01 |
| How frequently it is used (e.g. before sex, once a day, once a month) | 39 (10) | 35 (18) | 4 (2) | <0.01 |
| Side effects | 39 (10) | 31 (16) | 8 (4) | <0.01 |
| Ability to breastfeed and have healthy baby | 26 (7) | 15 (8) | 11 (5) | 0.32 |
| How it is used (e.g. vaginal ring, injected, pill) | 25 (6) | 16 (8) | 9 (4) | 0.10 |
| Privacy (from partner) | 13 (3) | 11 (6) | 2 (1) | 0.01 |
| Other (not forgotten easily, overall safety) | 2 (1) | 2 (1) | 0 (0) | 0.23 |
| **Most important access-related characteristic of a potential HIV prevention product (ranked #1)** |  |  |  |  |
| Free of charge | 203 (52) | 104 (55) | 99 (49) | 0.22 |
| How easy it is to obtain | 79 (20) | 43 (23) | 36 (18) | 0.22 |
| Where to pick up (clinic, pharmacy, community) | 48 (12) | 28 (15) | 20 (10) | 0.14 |
| Total time it takes to get product | 31 (8) | 15 (8) | 16 (8) | 0.99 |
| **Most preferable frequency of HIV prevention method (ranked #1)** |  |  |  |  |
| Before sex | 53 (13) | 30 (16) | 23 (11) | 0.19 |
| After sex | 6 (2) | 5 (3) | 1 (1) | 0.10 |
| Every day | 46 (12) | 26 (14) | 20 (10) | 0.23 |
| Once per month | 62 (16) | 25 (13) | 37 (18) | 0.18 |
| Once every 2-3 months | 59 (15) | 17 (9) | 42 (21) | <0.01 |
| Once every 6 months | 45 (11) | 11 (6) | 34 (17) | <0.01 |
| Once yearly | 123 (31) | 76 (40) | 47 (23) | <0.01 |

### Preference of long-acting PrEP modalities

Overall, three-fourths of participants (n=297, South Africa 74%, Kenya 76%, p=0.60) responded that they would prefer to switch to injectable PrEP over remaining on oral PrEP if it were available (**Figure 1**). South African and Kenyan women differed in their reasons for preferring injectable PrEP over oral PrEP: participants in South Africa more commonly preferred it for its longer duration of effectiveness (South Africa 87%, Kenya 42%, p<0.01) and not having to take a daily pill (South Africa 57%, Kenya 41%, p<0.01), while participants in Kenya were more likely to state reasons of privacy (Kenya 49%, South Africa 5%, p<0.01) and not having to carry pills (Kenya 42%, South Africa 13%, p<0.01) (**Supplementary Table 1**). The most common concerns regarding injectable PrEP were injection pain and potential side effects. Although overall these concerns were not frequently reported, Kenyan participants were more concerned than South African participants regarding the safety of the new injectable (Kenya 13%, South Africa 4%, p=0.01) and its safety for infants if using the injectable while breastfeeding (Kenya 6%, South Africa 0%, p=0.01).

**Figure 1:**
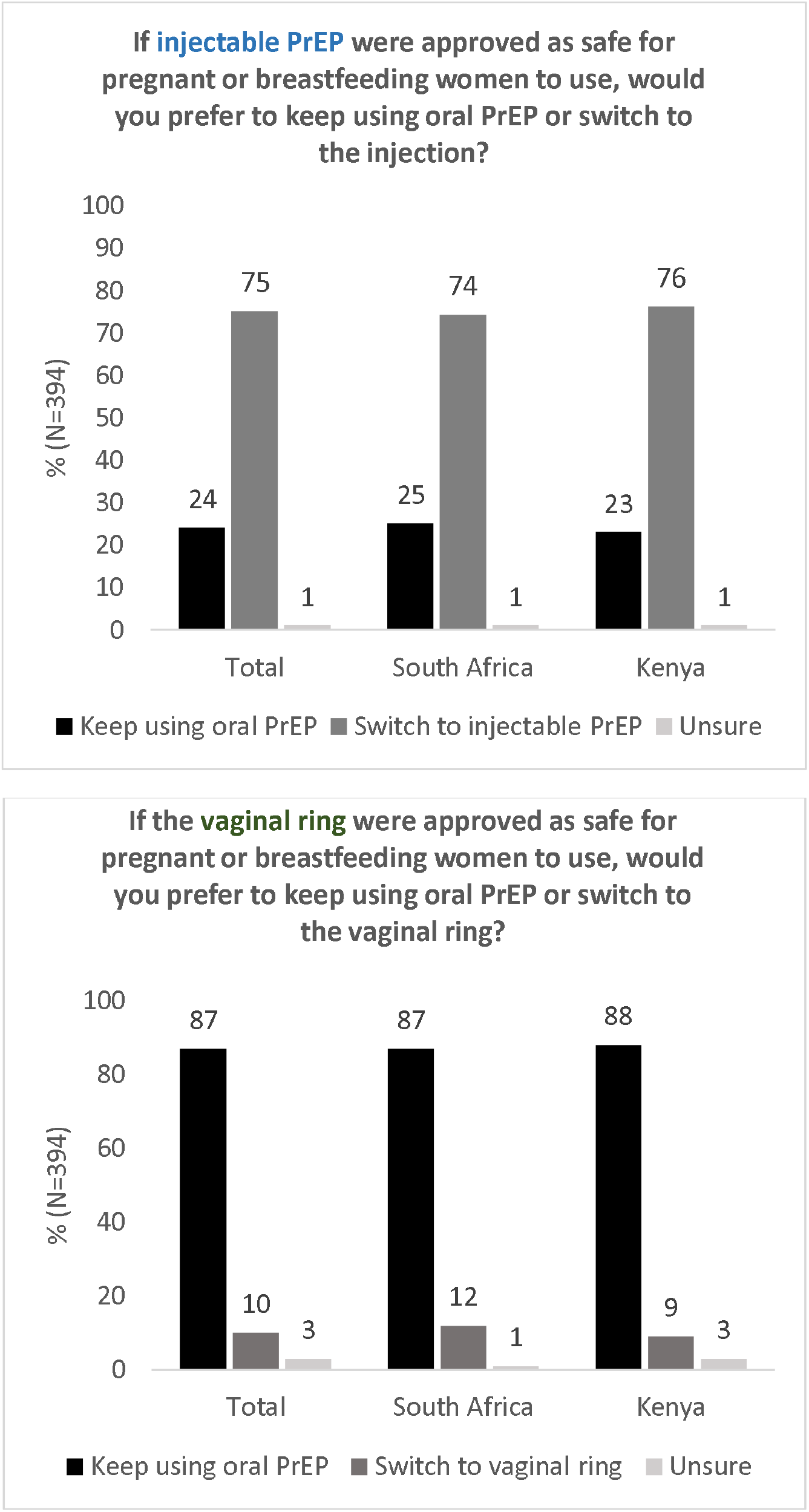
Theoretical acceptability of long-acting PrEP methods versus daily oral PrEP by pregnant and postpartum women with experience taking oral PrEP, Kenya and South Africa, September 2021 – February 2022 (N=394 women)

Fewer women (n=40, 10%) would prefer switching to the vaginal ring over oral PrEP in both South Africa and Kenya (12% and 9% respectively). South African women were interested in the vaginal ring most frequently due to its longer duration (n=14/22, 64%) and not having to remember to take a daily pill (n=12/22, 55%), while most common reasons for preferring the vaginal ring in Kenya were because it was “easy to use” (n=8/18, 44%) or did not involve carrying pills (n=6/18, 33%). Participants were unsure or preferred oral PrEP over the vaginal ring most frequently due to its insertion into the vagina particularly among South African women (South Africa 82%, Kenya 48%, p<0.01), potential side effects more so among Kenyan women (South Africa 21%, Kenya 33%, p=0.02), and concerns about safety (South Africa 23%, Kenya 27%). Participants in South Africa were also more frequently concerned about the vaginal ring not providing sufficient protection against HIV (14%, Kenya 7%, p=0.03).

In multivariable analyses adjusting for age and country, preference of a long-acting PrEP method (either injectable PrEP or vaginal ring) over daily oral PrEP was associated with prior use of injectable contraception (aOR=2.48, 95% CI=1.34, 4.57), disliking at least one attribute of daily oral PrEP (aOR=1.72, 95% CI=1.05, 2.80), disliking the daily use of oral PrEP (aOR=1.92, 95% CI=1.01, 3.67), and preferring longer duration of effectiveness (aOR=1.58, 95% CI=0.94, 2.65) (**Table 4**). Preferring a long-acting PrEP method over oral PrEP was negatively associated with previous use of the female condom (aOR=0.27, 95% CI=0.07, 1.12) and liking taking oral PrEP daily (aOR 0.30, 95% CI=0.07, 1.24).

**Table 4:** Factors associated with of long-acting PrEP preference vs. oral PrEP among pregnant and postpartum women with experience taking oral PrEP, N=394.

|  | Summary Statistics |  |  |  |  |  |
| --- | --- | --- | --- | --- | --- | --- |
|  | Wants long-acting<br>(n=305, %) | Wants oral PrEP<br>(n=89, %) | Unadjusted OR<br>(95% CI) | p-value | Adjusted OR<sup>†</sup><br>(95% CI) | p-value |
| *Maternal age (median, IQR) years* | 28 [24-32] | 27 [23-31] | 1.03 [0.99-1.08] | 0.11 | 1.04 [0.99, 1.08] | 0.11 |
| ≥25 years | 221 (72) | 59 (66) | 1.34 [0.81-2.22] | 0.26 | 1.35 [0.81-2.27] | 0.25 |
| <25 years | 84 (28) | 30 (34) |  |  |  |  |
| *Country* |  |  |  |  |  |  |
| Kenya | 158 (52) | 46 (52) | 1.01 [0.63-1.61] | 0.98 | 0.93 [0.57, 1.51] | 0.77 |
| South Africa | 147 (48) | 43 (48) |  |  |  |  |
| *Pregnancy status* |  |  |  |  |  |  |
| Pregnant | 78 (26) | 27 (30) | 0.79 [0.47-1.33] | 0.37 | 0.82 [0.48, 1.39] | 0.46 |
| Postpartum | 227 (74) | 62 (70) |  |  |  |  |
| *Time on PrEP (median, IQR) days* | 331 [168-422] | 336 [185-420] | 1.00 [1.00-1.00] | 0.90 | 1.00 [1.00-1.00] | 0.78 |
| *Last grade completed (ref: primary school)* | 27 (9) | 10 (11) | *ref* | *ref* | *ref* | *ref* |
| Some secondary | 178 (58) | 64 (72) | 1.20 [0.55-2.64] | 0.64 | 1.12 [0.52, 2.74] | 0.67 |
| Completed secondary | 72 (24) | 17 (19) | 1.48 [0.61-3.61] | 0.39 | 1.62 [0.61, 4.32] | 0.34 |
| Some or all tertiary | 20 (7) | 3 (3) | 1.78 [0.53-5.94] | 0.35 | 1.68 [0.49, 5.79] | 0.41 |
| *Currently employed (formally or informally)* |  |  |  |  |  |  |
| Employed | 61/304 (20) | 19 (21) | 0.93 [0.52-1.65] | 0.79 | 0.81 [0.44-1.49] | 0.49 |
| Not employed | 243/304 (80) | 70 (79) |  |  |  |  |
| *At least one sexual partner* |  |  |  |  |  |  |
| ≥1 sexual partner | 288 (94) | 85 (96) | 0.80 [0.26-2.43] | 0.69 | 0.75 [0.24-2.30] | 0.61 |
| 0 sexual partners | 16 (5) | 4 (4) |  |  |  |  |
| *PrEP stigma (median, IQR)* | 14 [13-17] | 15 [13-16] | 1.02 [0.96-1.09] | 0.56 | 1.03 [0.96-1.10] | 0.45 |
| Endorsed 0 stigma | 193/293 (66) | 58/87 (67) | 1.03 [0.62-1.72] | 0.89 | 1.06 [0.63-1.77] | 0.83 |
| Endorsed ≥1 stigma statement | 100/293 (34) | 29/87 (33) |  |  |  |  |
| *HIV Risk Perception* |  |  |  |  |  |  |
| Any perceived HIV risk | 194/291 (67) | 63/87 (72) | 0.76 [0.45, 1.29] | 0.31 | 0.75 [0.43, 1.31] | 0.31 |
| No perceived HIV risk | 97/291 (33) | 24/87 (28) |  |  |  |  |
| *Alcohol use* |  |  |  |  |  |  |
| Endorses any alcohol use | 12/293 (4) | 5/87 (6) | 0.70 [0.24-2.05] | 0.52 | 0.73 [0.25-2.17] | 0.58 |
| Does not endorse alcohol use | 281/293 (96) | 82/87 (94) |  |  |  |  |
| *PrEP persistence (has the participant taken PrEP over the past 30 days?)* |  |  |  |  |  |  |
| Yes | 222 (73) | 72 (81) | 0.63 [0.35-1.13] | 0.12 | 0.65 [0.36, 1.19] | 0.16 |
| No | 83 (27) | 17 (19) |  |  |  |  |
| *Time traveled to clinic and return (median, IQR)* | 40 [30-60] | 40 [30-60] | 1.00 [0.99-1.01] | 0.95 | 0.99 [0.99-1.01] | 0.99 |
| *minutes* |  |  |  |  |  |  |
| *Cost to travel to clinic and return (median, IQR)* | 1.29 [1.29-1.71] | 1.29 [1.29-1.71] | 1.12 [0.86-1.45] | 0.40 | 1.11 [0.85-1.45] | 0.44 |
| *USD equivalent* |  |  |  |  |  |  |
| *Contraceptive used in the past* |  |  |  |  |  |  |
| **Injection** | **252 (83)** | **60 (67)** | **2.30 [1.35-3.92]** | **0.002** | **2.48 [1.34, 4.57]** | **<0.01** |
| Condom | 172 (56) | 45 (51) | 1.26 [0.79-2.03] | 0.33 | 1.59 [0.82, 3.07] | 0.17 |
| Implant | 134 (44) | 47 (53) | 0.70 [0.44-1.12] | 0.14 | 0.71 [0.44, 1.16] | 0.17 |
| Oral contraceptive | 79 (26) | 19 (21) | 1.29 [0.73-2.27] | 0.38 | 1.22 [0.69-2.17] | 0.50 |
| **Female condom** | **4 (1)** | **4 (4)** | **0.28 [0.07-1.15]** | **0.08** | **0.27 [0.07-1.12]** | **0.07** |
| Vaginal ring | 1 (0.3) | 1 (0.3) | 0.29 [0.02-4.68] | 0.38 | 0.27 [0.02-4.42] | 0.36 |
| *Oral PrEP likes* |  |  |  |  |  |  |
| HIV prevention | 298 (98) | 87 (98) | 0.98 [0.20-4.80] | 0.98 | 0.95 [0.19-4.66] | 0.95 |
| No side effects | 50 (16) | 11 (12) | 1.39 [0.69-2.80] | 0.36 | 1.43 [0.71-2.90] | 0.32 |
| Ease of use | 21 (7) | 6 (7) | 1.02 [0.40-2.62] | 0.96 | 1.07 [0.41-2.75] | 0.90 |
| No interruption of sex | 12 (4) | 4 (4) | 0.87 [0.27-2.77] | 0.81 | 0.87 [0.27-2.80] | 0.82 |
| Easy to hide | 7 (2) | 3 (3) | 0.67 [0.17-2.66] | 0.57 | 0.63 [0.16-2.56] | 0.52 |
| **Take it daily** | **4 (1)** | **4 (4)** | **0.28 [0.07-1.15]** | **0.08** | **0.30 [0.07-1.24]** | **0.10** |
| Take it orally | 4 (1) | 3 (3) | 0.38 [0.08-1.73] | 0.21 | 0.38 [0.08-1.77] | 0.22 |
| Nothing | 1 (0.3) | 0 (0) | -- | -- | --- | --- |
| *Oral PrEP dislikes* |  |  |  |  |  |  |
| **Dislikes at least one thing about oral PrEP** | **158 (52)** | **35 (39)** | **1.66 [1.03-2.68]** | **0.04** | **1.72 [1.05, 2.80]** | **0.03** |
| Side effects | 81 (27) | 18 (20) | 1.43 [0.80-2.54] | 0.23 | 1.43 [0.80, 2.55] | 0.23 |
| **Daily use** | **75 (25)** | **13 (15)** | **1.91 [1.00-3.63]** | **0.05** | **1.92 [1.01, 3.67]** | **0.05** |
| Take it orally | 28 (9) | 7 (8) | 1.18 [0.50-2.81] | 0.70 | 1.20 [0.50, 2.85] | 0.69 |
| Not discreet | 7 (2) | 2 (2) | 1.02 [0.21-5.01] | 0.98 | 1.09 [0.22, 5.46] | 0.92 |
| No STI protection | 6 (2) | 4 (4) | 0.43 [0.12-1.55] | 0.20 | 0.45 [0.12, 1.64] | 0.23 |
| *Preferred frequency of PrEP use* |  |  |  |  |  |  |
| **Ranked every month, 2-3 months, 6 months, or year #1** | **230 (75)** | **59 (66)** | **1.56 [0.94-2.60]** | **0.09** | **1.58 [0.94, 2.65]** | **0.08** |
| Ranked before sex, after sex, or every day #1 | 75 (25) | 30 (34) |  |  |  |  |
| *Interested in community PrEP delivery* |  |  |  |  |  |  |
| Yes | 131 (43) | 32 (36) | 1.34 [0.82-2.19] | 0.24 | 1.45 [0.85, 2.45] | 0.17 |
| No | 174 (57) | 57 (64) |  |  |  |  |

† Each individual model adjusted for maternal age, country **Bold $p < 0.10$**

## Discussion

We identified a strong theoretical preference for long-acting injectable PrEP among pregnant and postpartum women in Kenya and South Africa. Additionally, about one-half of women reported liking daily oral PrEP, indicating the importance of providing pregnant and postpartum women with choices of modalities. Studies assessing end-user preference among African women of potential HIV prevention methods (multiple vaginally inserted methods, injection, pill) showed that women had varied preference for these HIV prevention modalities, and women described the importance of personal preference in product choice[32-34]. Similar to HIV prevention, studies on contraceptive choice indicate that the best method for an individual depends on their preferences, necessitating a diversity of contraceptive options[35-37]. Our data suggest that the availability of choices to meet personal preferences is essential to improving overall uptake of HIV prevention methods among pregnant and postpartum women as well. Furthermore, women in South Africa and Kenya did not differ in preferences for long-acting PrEP modalities compared to oral PrEP, but did differ in the reasoning behind their preferences, indicating the importance of context-specific implementation when providing HIV prevention modalities. Sociodemographic characteristics and available PrEP modalities differ between countries, indicating that differences between countries in PrEP preferences among pregnant and postpartum women need to be studied and incorporated into country-specific plans for PrEP rollout.

Our findings on preference for long-acting injectable PrEP among pregnant and postpartum women, as well as most reasons for their PrEP preferences, align with the existing literature among nonpregnant women. Women also noted need for safety data in pregnancy and lactation in our study, which was distinct for this population. In studies providing hypothetical choices between long-acting PrEP modalities, nonpregnant women from Kenya and South Africa preferred injectable methods of HIV prevention over other modalities, despite similar concerns regarding injection pain[38-40]. In studies assessing acceptability of long-acting injectable PrEP among women within clinical trials in Africa and the United States, participants described acceptability of long-acting injectable PrEP due to its ease of use and long-term protection and would use the method again[41-43], which may suggest that hypothetical acceptability of long-acting injectable PrEP among pregnant and postpartum women may align with its acceptability among nonpregnant women during implementation. Some PrEP preferences identified by women in our study were specific to the pregnancy-postpartum period, such as the importance of safety in pregnancy and infant safety while breastfeeding on PrEP. Previous data have identified the positive impacts of counseling about infant safety on oral PrEP initiation among women in Kenya, highlighting that obtaining further data on pregnancy outcomes and infant safety of CAB-LA, as well as counseling on safety may be beneficial for uptake[18].

Pregnant and postpartum women in our study reported that they would prefer to use a long-acting injectable PrEP for privacy and discreetness of this method, highlighting the role of long-acting PrEP to mitigate sociocultural barriers previously identified with oral PrEP use[16-18,44]. Qualitative studies in Kenya and South Africa have described the negative community perception surrounding PrEP use experienced by pregnant and nonpregnant women, particularly due to its association with high-risk sexual behavior as well as being conflated with antiretroviral therapy, leading to concealment of product use[45-47]. A qualitative study assessing oral PrEP perceptions among pregnant and postpartum women in Kenya additionally described fear of male partners becoming violent if discovering their PrEP use, indicating that stigma and negative sentiments towards PrEP within a household may decrease PrEP disclosure and necessitate concealment[48,49]. The availability of a HIV prevention method, such as long-acting injectable PrEP, that does not require daily administration or concealment of physical pills may make it easier for pregnant and postpartum women experiencing PrEP stigma to persist on the HIV prevention method – and has been hypothetically posed by women as a potential solution in previous qualitative work[47]. Long-acting PrEP modalities must be made available in conjunction to other solutions that mitigate community- and individual-level stigma experienced by women in Kenya and South Africa, such as community-facing interventions involving media and educational initiatives, as well as the involving of male partners in HIV prevention and education[41,50].

Few pregnant and postpartum women reported preference for the PrEP vaginal ring over oral PrEP, which may be due to unfamiliarity with the method as well as its lower efficacy. Existing acceptability and demonstration studies of the dapivirine ring for HIV prevention show that despite similar initial concerns regarding insertion into the vagina and potential side effects, women in Sub-Saharan Africa who began using vaginal rings for HIV prevention developed familiarity with the method, found it easy to integrate into their lives, and reported willingness to use the method in the future[51-54]. Our data in the context of these studies indicate the importance of education regarding vaginal ring insertion and anticipated side effects when counseling about available HIV prevention modalities, and reemphasize the need for available choice in HIV prevention methods that can be used individually or in combination.

Prior use of injectable contraception was associated with a preference for injectable PrEP, indicating that contraceptive knowledge and familiarity with the concept of regular injections as a prevention measure may impact uptake of injectable PrEP among pregnant and postpartum women. Studies assessing acceptability of multipurpose prevention technologies combining HIV and pregnancy prevention similarly showed that injections were preferred compared to a ring or pill-form of combination prevention, and that past experience with similar contraceptive delivery forms was a significant predictor of their method choice[55-57]. Combining HIV prevention education with family planning education may improve uptake of HIV prevention, due to existing familiarity with long-acting contraceptive modalities. Further, the future development of combination prevention methods may improve uptake of HIV prevention.

### Limitations

Our study included pregnant and postpartum women with knowledge and experience with oral PrEP. As such, our results may differ from preferences of pregnant and postpartum women without prior experience using PrEP. Our surveys provided a hypothetical choice between PrEP modalities, as some methods are not yet available in South Africa and Kenya (i.e. injectable PrEP in both countries, vaginal ring PrEP in Kenya). Responses may differ from PrEP choices during an implementation offering multiple modalities. Our results may not be generalizable to other settings beyond urban South Africa and Kenya, so further study is necessary on the acceptability of long-acting PrEP among pregnant and postpartum women in other settings.

## Conclusions

Our study demonstrates that pregnant and postpartum women on oral PrEP may desire a switch to long-acting injectable PrEP as it becomes available in South Africa and Kenya. Women in our study had different PrEP preferences and different reasons for these preferences, indicating the need for client-centered PrEP programs that offer diverse choices for HIV prevention and empower women to make the choice best suited to their needs and lifestyle. Some of these preferences, such as safety for maternal-infant dyad, were specific to the pregnant-postpartum period, while others likely reflected general preferences of women about PrEP. Overall, further efforts are needed to increase choice and accessibility of various PrEP options to benefit the well-being of pregnant and postpartum women and their infants.

## Supporting information

COI Disclosure

STROBE Checklist

## Data Availability

All data produced in the present study are available upon reasonable request to the corresponding author, Nafisa Wara.

## Authorship

Study was conceived by NJW and designed by NJW, RH, JP, DLJD. NJW oversaw study implementation and data collection, analyzed the data, wrote the first draft of manuscript, and reviewed the manuscript following revision by all co-authors. RM, MM, LG, and NM reviewed the study design, oversaw study implementation and data collection, and assisted with data analysis. CO, CM, RH, JP, and DLJD reviewed the study design, study data, and data analysis. JK, GJ-S, and JP designed the PrIMA-X study, and LM and DLJD designed the PrEP-PP study. All authors contributed to the development of this manuscript, and have read and approved the final manuscript.

## Acknowledgments

We would like to thank all PrEP-PP and PrIMA-X study participants for their time and perspectives, as well as the PrEP-PP and PrIMA-X project staff, the City of Cape Town Department of Health, Kenyan Ministry of Health, and Siaya and Homa Bay County Governments.

---

**Supplementary Table 1:**
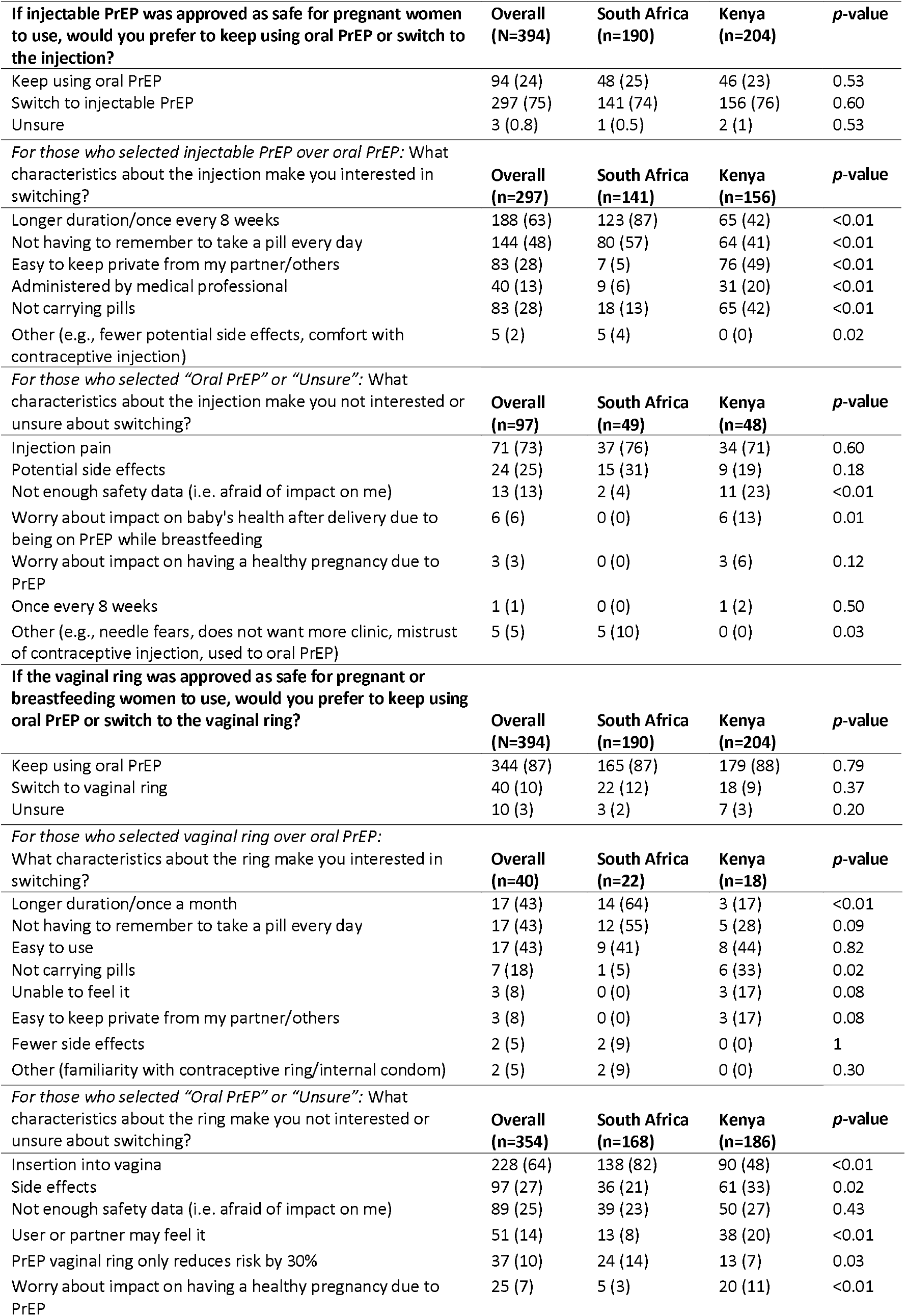
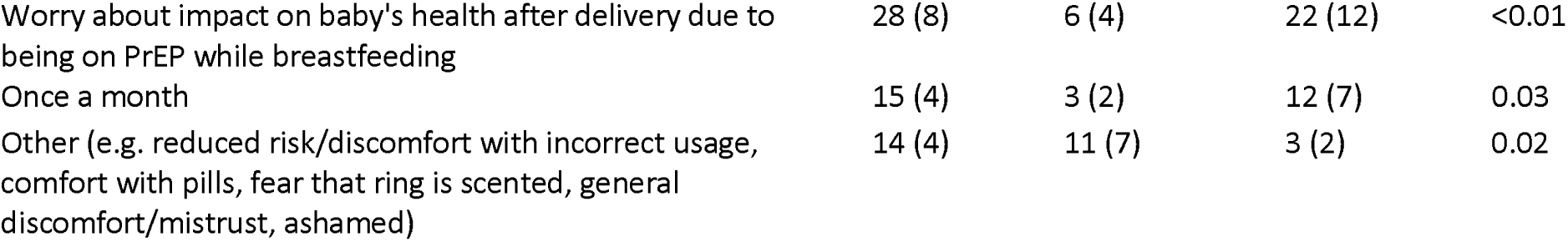
Reasons behind preference of long-acting PrEP methods versus oral PrEP methods by pregnant and postpartum women with experience taking oral PrEP, South Africa and Kenya.

| **If injectable PrEP was approved as safe for pregnant women to use, would you prefer to keep using oral PrEP or switch to the injection?** | **Overall (N=394)** | **South Africa (n=190)** | **Kenya (n=204)** | **p-value** |
| --- | --- | --- | --- | --- |
| Keep using oral PrEP | 94 (24) | 48 (25) | 46 (23) | 0.53 |
| Switch to injectable PrEP | 297 (75) | 141 (74) | 156 (76) | 0.60 |
| Unsure | 3 (0.8) | 1 (0.5) | 2 (1) | 0.53 |
| *For those who selected injectable PrEP over oral PrEP: What characteristics about the injection make you interested in switching?* |  |  |  |  |
|  | **Overall (n=297)** | **South Africa (n=141)** | **Kenya (n=156)** | **p-value** |
| Longer duration/once every 8 weeks | 188 (63) | 123 (87) | 65 (42) | <0.01 |
| Not having to remember to take a pill every day | 144 (48) | 80 (57) | 64 (41) | <0.01 |
| Easy to keep private from my partner/others | 83 (28) | 7 (5) | 76 (49) | <0.01 |
| Administered by medical professional | 40 (13) | 9 (6) | 31 (20) | <0.01 |
| Not carrying pills | 83 (28) | 18 (13) | 65 (42) | <0.01 |
| Other (e.g., fewer potential side effects, comfort with contraceptive injection) | 5 (2) | 5 (4) | 0 (0) | 0.02 |
| *For those who selected "Oral PrEP" or "Unsure": What characteristics about the injection make you not interested or unsure about switching?* |  |  |  |  |
|  | **Overall (n=97)** | **South Africa (n=49)** | **Kenya (n=48)** | **p-value** |
| Injection pain | 71 (73) | 37 (76) | 34 (71) | 0.60 |
| Potential side effects | 24 (25) | 15 (31) | 9 (19) | 0.18 |
| Not enough safety data (i.e. afraid of impact on me) | 13 (13) | 2 (4) | 11 (23) | <0.01 |
| Worry about impact on baby's health after delivery due to being on PrEP while breastfeeding | 6 (6) | 0 (0) | 6 (13) | 0.01 |
| Worry about impact on having a healthy pregnancy due to PrEP | 3 (3) | 0 (0) | 3 (6) | 0.12 |
| Once every 8 weeks | 1 (1) | 0 (0) | 1 (2) | 0.50 |
| Other (e.g., needle fears, does not want more clinic, mistrust of contraceptive injection, used to oral PrEP) | 5 (5) | 5 (10) | 0 (0) | 0.03 |
| **If the vaginal ring was approved as safe for pregnant or breastfeeding women to use, would you prefer to keep using oral PrEP or switch to the vaginal ring?** |  |  |  |  |
|  | **Overall (N=394)** | **South Africa (n=190)** | **Kenya (n=204)** | **p-value** |
| Keep using oral PrEP | 344 (87) | 165 (87) | 179 (88) | 0.79 |
| Switch to vaginal ring | 40 (10) | 22 (12) | 18 (9) | 0.37 |
| Unsure | 10 (3) | 3 (2) | 7 (3) | 0.20 |
| *For those who selected vaginal ring over oral PrEP: What characteristics about the ring make you interested in switching?* |  |  |  |  |
|  | **Overall (n=40)** | **South Africa (n=22)** | **Kenya (n=18)** | **p-value** |
| Longer duration/once a month | 17 (43) | 14 (64) | 3 (17) | <0.01 |
| Not having to remember to take a pill every day | 17 (43) | 12 (55) | 5 (28) | 0.09 |
| Easy to use | 17 (43) | 9 (41) | 8 (44) | 0.82 |
| Not carrying pills | 7 (18) | 1 (5) | 6 (33) | 0.02 |
| Unable to feel it | 3 (8) | 0 (0) | 3 (17) | 0.08 |
| Easy to keep private from my partner/others | 3 (8) | 0 (0) | 3 (17) | 0.08 |
| Fewer side effects | 2 (5) | 2 (9) | 0 (0) | 1 |
| Other (familiarity with contraceptive ring/internal condom) | 2 (5) | 2 (9) | 0 (0) | 0.30 |
| *For those who selected "Oral PrEP" or "Unsure": What characteristics about the ring make you not interested or unsure about switching?* |  |  |  |  |
|  | **Overall (n=354)** | **South Africa (n=168)** | **Kenya (n=186)** | **p-value** |
| Insertion into vagina | 228 (64) | 138 (82) | 90 (48) | <0.01 |
| Side effects | 97 (27) | 36 (21) | 61 (33) | 0.02 |
| Not enough safety data (i.e. afraid of impact on me) | 89 (25) | 39 (23) | 50 (27) | 0.43 |
| User or partner may feel it | 51 (14) | 13 (8) | 38 (20) | <0.01 |
| PrEP vaginal ring only reduces risk by 30% | 37 (10) | 24 (14) | 13 (7) | 0.03 |
| Worry about impact on having a healthy pregnancy due to PrEP | 25 (7) | 5 (3) | 20 (11) | <0.01 |
| Worry about impact on baby's health after delivery due to being on PrEP while breastfeeding | 28 (8) | 6 (4) | 22 (12) | <0.01 |
| Once a month | 15 (4) | 3 (2) | 12 (7) | 0.03 |
| Other (e.g. reduced risk/discomfort with incorrect usage, comfort with pills, fear that ring is scented, general discomfort/mistrust, ashamed) | 14 (4) | 11 (7) | 3 (2) | 0.02 |

